# Association between inflammatory bowel disease and prostate cancer: A large-scale, prospective, population-based study

**DOI:** 10.1101/2020.01.16.20017707

**Authors:** Travis J. Meyers, Adam B. Weiner, Rebecca E. Graff, Anuj S. Desai, Lauren Folgosa Cooley, William J. Catalona, Stephen B. Hanauer, Jennifer D. Wu, Edward M. Schaeffer, Sarki A. Abdulkadir, Shilajit D. Kundu, John S. Witte

**Affiliations:** Department of Epidemiology and Biostatistics, University of California, San Francisco (UCSF), San Francisco, CA; Department of Urology, Northwestern University Feinberg School of Medicine, Chicago, IL; Department of Medicine, Division of Gastroenterology and Hepatology, Northwestern University Feinberg School of Medicine, Chicago, IL; Department of Urology, UCSF

**Keywords:** cohort study, epidemiology, inflammatory bowel disease, prostate cancer, UK Biobank, ulcerative colitis

## Abstract

**Background:** Inflammatory bowel disease (IBD) is an established risk factor for colorectal cancer. Recent reports suggesting IBD is also a risk factor for prostate cancer (PC) require further investigation.

**Objective:** To test the association between IBD with incident PC.

**Design, setting, and participants:** We studied 218,084 men in the population-based UK Biobank cohort, aged 40-69 at study entry between 2006 and 2010, with follow-up through mid-2015.

**Outcome measurements and statistical analysis:** We assessed the association between IBD and subsequent PC using multivariable Cox regression analyses, adjusting for age at assessment, ethnic group, UK region, smoking status, alcohol drinking frequency, body mass index, Townsend Deprivation Index, family history of prostate cancer, and previous prostate-specific antigen testing.

**Results and limitations:** Mean age at study entry was 56 years, 94% of the men were white, and 1.1% (n=2,311) had a diagnosis of IBD. After a median follow-up of 78 months, men with IBD had an increased risk of PC (adjusted hazards ratio [aHR] = 1.31, 95% Confidence interval [CI] = 1.03-1.67, p = 0.029). Separately analyzing the IBD subtypes of ulcerative colitis (UC) and Crohn’s disease (CD), the association with PC was only among men with the former (UC; aHR = 1.47, 95% CI = 1.11-1.95, p=0.0070), and not the latter (CD; aHR 1.06, 95% CI = 0.63-1.80, p = 0.82). Results are limited by lack of data on frequency of health care interactions.

**Conclusions:** In a large-scale, prospective cohort study, we detected an association between IBD, and UC specifically, with incident PC diagnosis.

**Patient summary:** This study of over 200,000 men in the UK suggests that men with inflammatory bowel disease may be at a higher risk of prostate cancer than the general population.

## Introduction

Prostate cancer (PC) is the second most common non-cutaneous malignancy in men globally, accounting for 1.3 million new cases and over 350,00 deaths in 2018 globally^1^. Screening for PC may help reduce PC mortality at the potential cost of overdiagnosis leading to unnecessary exposure to treatment related morbidities^2,3^. Guidelines in both the United States and Europe acknowledge the benefits of identifying risk factors for PC to better counsel men on the use of prostate-specific antigen (PSA)- based screening^4,5^.

While inflammatory bowel disease (IBD) is an established risk factor for colorectal cancer^6^, associations between IBD with prostate cancer have been reported for some studies^7–13^ but not others^14–16^. We thus conducted a prospective study of men from the large-scale, population-based UK Biobank cohort^17^ to test this association. Men in the UK historically have low rates of PC screening (3-11% after age 50 years)^18^ and PC screening is not currently recommended by the UK National Screening Committee^19^. We hypothesized that men in the UK Biobank with a diagnosis of IBD would experience a higher incidence of subsequent PC diagnosis.

## Patients and Methods

### Study population

The UK Biobank is a prospective, population-based study established to investigate genetic and non-genetic risk factors for disease in individuals of middle and advanced age^17^. The details of study design and data collection have previously been described and the complete protocol can be found online^17,20^. In summary, 500,796 participants aged 40-69 years registered within the National Health Service (NHS) and living within 40km of one of 22 assessment centers across England, Scotland, and Wales were recruited between 2006 and 2010. Each participant provided written informed consent and the UK Biobank’s study protocol was approved by the UK North West Multicenter Research Ethics Committee. Baseline assessments at entry into the cohort were made for each participant in 90-minute appointments which included questionnaires, sample collections, and health care physical exams and interviews. Participants are linked to the NHS Central register to capture IBD status, cancer diagnoses, and deaths.

Since the present outcome of interest was prostate cancer, the analytical cohort was limited to self-reported male participants (n=228,284). We excluded men if at baseline assessment they had: 1) prior history of a malignant cancer (any site), or timing of malignant cancer diagnosis relative to baseline could not be determined (n=9,902, 4.3%); 2) surgical removal of the prostate (n=64, <0.01%; NHS procedure codes: OPCS version 3-630-635; OPCS version 4-M61); 3) earlier recorded death date (n=1, <0.01%). We also excluded 233 (<0.01%) individuals whose genetically inferred sex was female. The remaining 218,084 men comprised the study population.

### Exposure

The exposure of interest was a history of IBD (ulcerative colitis [UC] or Crohn’s disease [CD]) at the time of baseline assessment. IBD history was considered present if the participant had either a relevant inpatient ICD code or self-reported illness. ICD10 codes for UC and CD were K51 and K50, respectively. ICD9 codes for UC and CD were 556 and 555. Self-reported IBD and the approximate date that a doctor first diagnosed IBD were collected during the baseline assessment interview. If a participant had recorded diagnoses for both UC and CD, then they were still included in the analysis of overall IBD but not in the subtype analyses.

### Outcome

The outcome of interest was first diagnosis of malignant PC following baseline assessment. Prostate cancer case status was determined using ICD codes (ICD-9: 185, ICD-10: C61). Follow-up data for the UK Biobank cohort was available through the middle of 2015.

### Covariates

Covariates included in multivariable analyses included age at baseline assessment (continuous), self-reported ethnicity (White, Mixed, South Asian, Chinese, Black, or other), region of assessment center (10 cancer registry regions), Townsend Deprivation Index (TDI; quintiles), smoking status (never, former, or current), alcohol drinking frequency (never, special occasions only, 1-3 times per month, 1-2 times per week, 3-4 times per week, daily/almost daily), body mass index (BMI; quintiles), family history of PC in biological relatives (yes, no), and history of PSA testing (yes, no). All categorical variables included a category for missingness.

### Statistical analyses

Person-years were calculated from the date of baseline assessment until diagnosis of PC, diagnosis of a different malignant cancer, prostatectomy, death, or end of follow-up, whichever came first. The non-PC endpoints were considered censoring events. If PC diagnosis followed any of the other censoring events within three months, then PC was used as the endpoint and the time to PC was included as the follow-up time. The rationale was that the censoring event may have been correlated with PC diagnosis.

Incidence rates for PC were calculated for each baseline group (no IBD, any IBD, UC, or CD) from the number of incident PC cases divided by the person-years of follow-up in each group. Kaplan-Meier curves were fit comparing survival for men with IBD (any IBD, UC, or CD) at baseline to those without IBD at baseline. The difference in survival by IBD status was compared by the univariate log-rank test.

Adjusted hazard ratios (aHRs) and 95% confidence intervals (CIs) for the association between IBD and PC were calculated using Cox proportional hazards models adjusting for all aforementioned covariates. This analysis was the basis for testing our main hypothesis. We further investigated whether the potential association between IBD and PC varied by: 1) IBD subtype; 2) duration of living with IBD (using 20 years from IBD diagnosis until baseline assessment in UK Biobank as the cut point); and 3) age at assessment (using 60 years as the cut point). For analysis 1 and 2, we performed the same regression as the primary analysis except IBD status was stratified as UC, CD, or no IBD and IBD > 20 year, IBD ≤ 20 years, or no IBD, respectively. For analysis 3, we created a new model which included age categorized as ≤60 or > 60 years as opposed to continuous and an interaction term between this new age variable and IBD status (IBD or no IBD).

To determine the appropriateness of our Cox models, we tested the underlying assumption that the relative incidence of PC between men with and without IBD was constant over time (proportional hazards). Specifically, the correlation between scaled Schoenfeld residuals with follow-up time was tested, based on the univariable Cox model with IBD status as the regressor. All statistical analyses were conducted using R statistical software, version 3.6.0: Kaplan-Meier plots were generated by the ‘survminer’ package; Cox models were conducted using the ‘survival’ package.

## Results

### Study participants

At baseline assessment, there were 2,311 men with a history of IBD and 215,773 men without a history of IBD (**Table 1**). Compared to men without IBD, men with IBD were on average one year older (57.3 vs. 56.5; p < 0.05), were more likely to be White (95.3% vs. 93.8%; p < 0.05), were more likely to be former smokers (49.5% vs. 37.7%) than current smokers (8.9% vs. 12.6%), and had a lower average BMI (27.5 vs. 27.8; p < 0.05). All participants were similar with respect to average TDI (No IBD: −1.25; IBD: - 1.19; negative values reflect relative affluence), family history of PC (both 7.7%), and history of PSA testing (No IBD: 27.6%; IBD: 26.9%). Of those with IBD, 1,488 exclusively had UC and 643 exclusively had CD. Compared to men with CD, men with UC were on average one year older (57.7 vs. 56.4; p < 0.05), had a lower TDI (−1.33 vs. −0.88; p < 0.05), were more likely to be former smokers (51.2% vs. 47.6%) than current smokers (6.7% vs. 13.7%), and had a higher average BMI (27.7 vs. 27.1, p < 0.05).

**Table 1.**
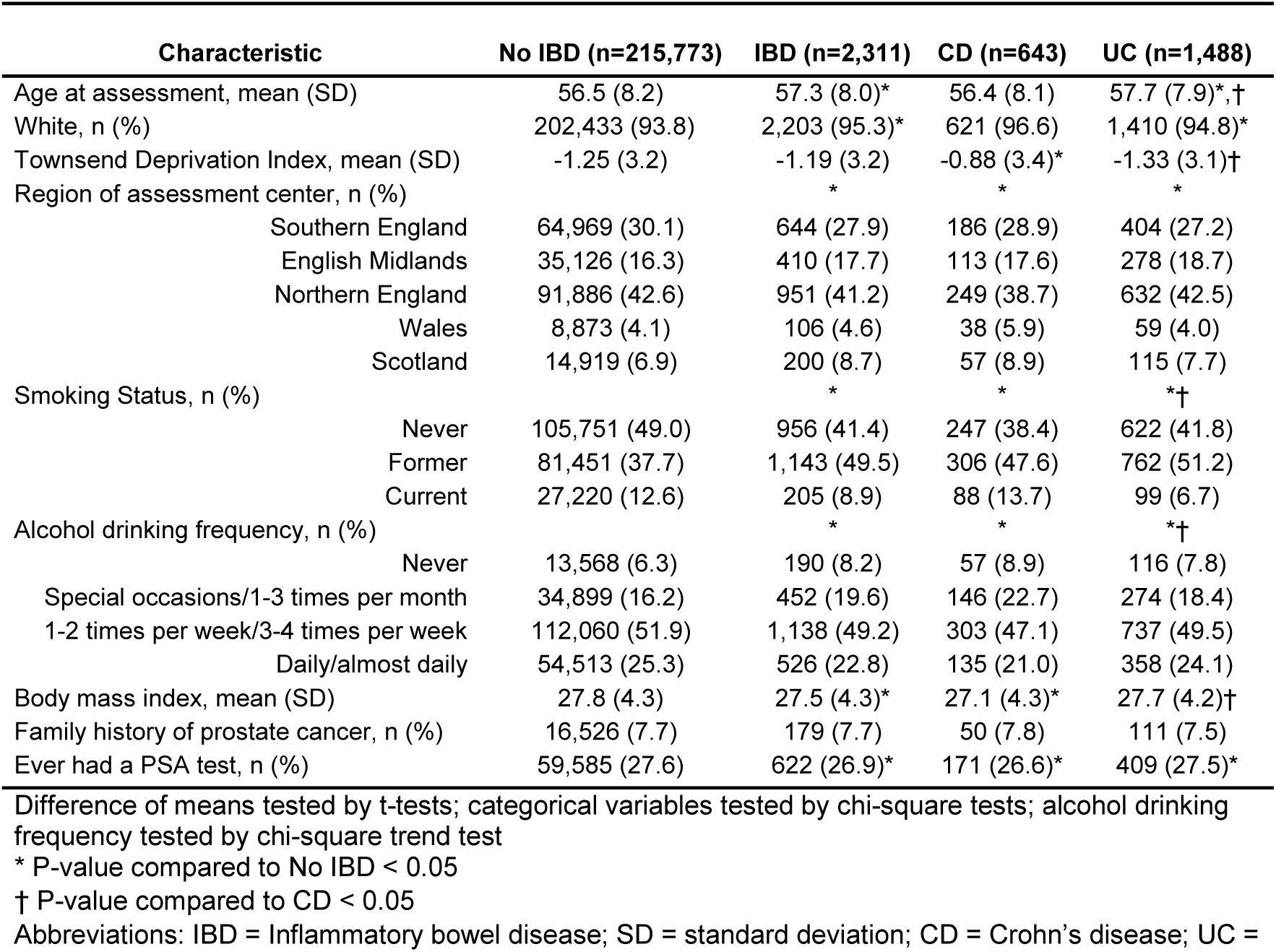
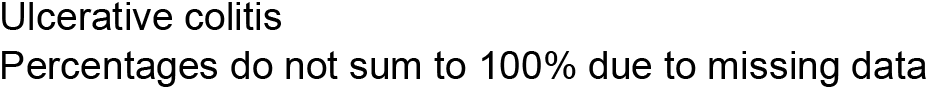
Baseline characteristics of male UK Biobank participants by IBD status at baseline assessment

| Characteristic | No IBD (n=215,773) | IBD (n=2,311) | CD (n=643) | UC (n=1,488) |
| --- | --- | --- | --- | --- |
| Age at assessment, mean (SD) | 56.5 (8.2) | 57.3 (8.0)* | 56.4 (8.1) | 57.7 (7.9)*,† |
| White, n (%) | 202,433 (93.8) | 2,203 (95.3)* | 621 (96.6) | 1,410 (94.8)* |
| Townsend Deprivation Index, mean (SD) | -1.25 (3.2) | -1.19 (3.2) | -0.88 (3.4)* | -1.33 (3.1)† |
| Region of assessment center, n (%) |  | * | * | * |
| Southern England | 64,969 (30.1) | 644 (27.9) | 186 (28.9) | 404 (27.2) |
| English Midlands | 35,126 (16.3) | 410 (17.7) | 113 (17.6) | 278 (18.7) |
| Northern England | 91,886 (42.6) | 951 (41.2) | 249 (38.7) | 632 (42.5) |
| Wales | 8,873 (4.1) | 106 (4.6) | 38 (5.9) | 59 (4.0) |
| Scotland | 14,919 (6.9) | 200 (8.7) | 57 (8.9) | 115 (7.7) |
| Smoking Status, n (%) |  | * | * | *† |
| Never | 105,751 (49.0) | 956 (41.4) | 247 (38.4) | 622 (41.8) |
| Former | 81,451 (37.7) | 1,143 (49.5) | 306 (47.6) | 762 (51.2) |
| Current | 27,220 (12.6) | 205 (8.9) | 88 (13.7) | 99 (6.7) |
| Alcohol drinking frequency, n (%) |  | * | * | *† |
| Never | 13,568 (6.3) | 190 (8.2) | 57 (8.9) | 116 (7.8) |
| Special occasions/1-3 times per month | 34,899 (16.2) | 452 (19.6) | 146 (22.7) | 274 (18.4) |
| 1-2 times per week/3-4 times per week | 112,060 (51.9) | 1,138 (49.2) | 303 (47.1) | 737 (49.5) |
| Daily/almost daily | 54,513 (25.3) | 526 (22.8) | 135 (21.0) | 358 (24.1) |
| Body mass index, mean (SD) | 27.8 (4.3) | 27.5 (4.3)* | 27.1 (4.3)* | 27.7 (4.2)† |
| Family history of prostate cancer, n (%) | 16,526 (7.7) | 179 (7.7) | 50 (7.8) | 111 (7.5) |
| Ever had a PSA test, n (%) | 59,585 (27.6) | 622 (26.9)* | 171 (26.6)* | 409 (27.5)* |

Difference of means tested by t-tests; categorical variables tested by chi-square tests; alcohol drinking frequency tested by chi-square trend test
\* P-value compared to No IBD < 0.05
† P-value compared to CD < 0.05
Abbreviations: IBD = Inflammatory bowel disease; SD = standard deviation; CD = Crohn's disease; UC =
Ulcerative colitis
Percentages do not sum to 100% due to missing data

### Prostate cancer incidence based on inflammatory bowel disease status

After a median follow-up of 78 months (over 1.3 million person-years for men without IBD and 14,379 years for men with IBD), there were 4,681 new cases of PC in men without IBD and 66 in men with IBD (**Table 2**). Men with IBD demonstrated a shorter time to developing PC (Log-rank, p=0.018; **Figure 1**). The assumption of proportional hazards was satisfied for each univariate model for baseline status of any IBD, UC, or CD. The incidence rates for PC (cases per 100,000 person-years) were 343 for non-IBD and 459 for men with IBD. After adjusting for covariates, IBD was associated with an increased hazard of PC (aHR = 1.31, 95% CI = 1.03 - 1.67, p = 0.029; **Table 2**). Further adjusting for history of partial or complete colectomy did not meaningfully change the HR so it was not included in the final models. In addition, we observed a trend for increasing HR across years since IBD diagnosis (<=20 years, aHR = 1.22; >20 years, aHR = 1.49; p-trend = 0.018; **Table 3)**. We did not detect interaction by age at study assessment (p=0.92).

**Table 2.** Cox regressions assessing the association between inflammatory bowel disease and future prostate cancer.

| **IBD status<sup>1</sup>** | **n** | **Person-years** | **PC cases** | **Incidence/100,000 PYs** | **Adjusted hazard ratio (95% CI)<sup>1</sup>** | **P** |
| --- | --- | --- | --- | --- | --- | --- |
| No IBD | 215,773 | 1,365,610 | 4,681 | 343 | Reference |  |
| Any IBD | 2,311 | 14,379 | 66 | 459 | 1.31 (1.03-1.67) | 0.029 |
| UC | 1,488 | 9,201 | 49 | 533 | 1.47 (1.11-1.95) | 0.0070 |
| CD | 643 | 4,021 | 14 | 348 | 1.06 (0.63-1.80) | 0.82 |

1. Three separate models were tested: Any IBD vs. No IBD; UC vs. No IBD; and CD vs. No IBD
2. Models adjusted for age at assessment, ethnic group, UK region, smoking status, alcohol drinking frequency, body mass index, Townsend Deprivation Index, family history of prostate cancer, and ever had a PSA test.
Abbreviations: IBD = Inflammatory bowel disease; UC = Ulcerative colitis; CD = Crohn's disease; PC = Prostate cancer; CI = Confidence interval; PYs = Person-years

**Table 3.** Adjusted hazard ratios and 95% CIs of IBD duration (diagnosis until assessment) and future PC, compared to never having IBD

| IBD Status<sup>1</sup> | <=20 years |  |  |  | >20 years |  |  |  | P for trend |
| --- | --- | --- | --- | --- | --- | --- | --- | --- | --- |
|  | n | Person-Years | PC Cases | HR (95% CI)<sup>2</sup> | n | Person-Years | PC Cases | HR (95% CI)<sup>2</sup> |  |
| Any IBD | 1,601 | 9,990 | 40 | 1.22<br>(0.89, 1.66)<br>p = 0.22 | 710 | 4,389 | 26 | 1.49<br>(1.01, 2.19)<br>p = 0.042 | 0.018 |
| UC | 1,076 | 6,679 | 29 | 1.29<br>(0.89, 1.85)<br>p = 0.18 | 412 | 2,521 | 20 | 1.87<br>(1.21, 2.91)<br>p = 0.0052 | 0.0022 |
| CD | 406 | 2,545 | 9 | 1.11<br>(0.58, 2.14)<br>p = 0.75 | 237 | 1,476 | 5 | 0.98<br>(0.41, 2.37)<br>p = 0.97 | 0.89 |

1. Three separate models were tested: Any IBD vs. No IBD; UC vs. No IBD; and CD vs. No IBD; No IBD: N= 215,773; 1.37 million person-years of follow-up; 4,681 PC cases
2. Models adjusted for age at assessment, ethnic group, UK region, smoking status, alcohol drinking frequency, body mass index, Townsend Deprivation Index, family history of prostate cancer, and ever had a PSA test
Abbreviations: IBD = Inflammatory bowel disease; UC = Ulcerative colitis; CD = Crohn's disease; PC = Prostate cancer; CI = Confidence interval

**Figure 1.**
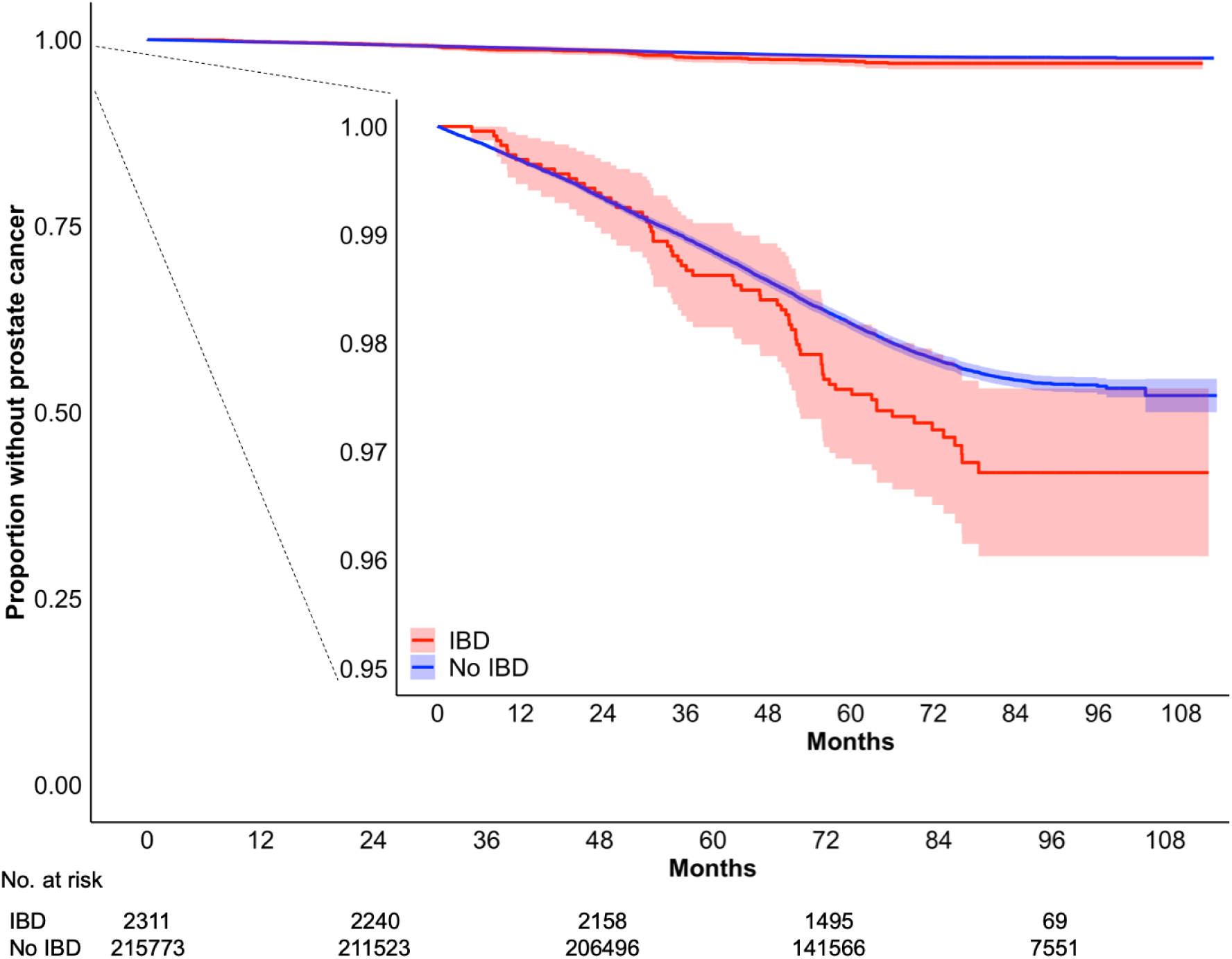
Kaplan-Meier analysis of prostate cancer incidence by inflammatory bowel disease status. Shading represents 95% confidence intervals. Log-rank comparing IBD to no IBD p=0.018. Abbreviations: IBD = Inflammatory bowel disease

### Prostate cancer incidence based on inflammatory bowel disease subtype

The person-years for men with exclusively one IBD subtype were comprised of 9,201 for men with UC and 4,021 for men with CD (**Table 2**). A total of 49 men with UC and 14 men with CD developed PC. While men with UC developed PC more rapidly than men without IBD (Log-rank p=0.0023, 533 cases per 100,000 person-years), those with CD did not (Log-rank p=0.95; 348 cases per 100,00 person-years; **Figure 2**). The same associations were noted on adjusted analysis (UC: aHR = 1.47, 95% CI = 1.11 - 1.95, p= 0.0070; CD: aHR = 1.06, 95% CI = 0.63 - 1.80, p = 0.82; **Table 2**). For the UC subtype, increasing HR was also noted across years since diagnosis (<=20 years, aHR = 1.29; >20 years, aHR = 1.87; p-trend = 0.0022; **Table 3**).

**Figure 2.**
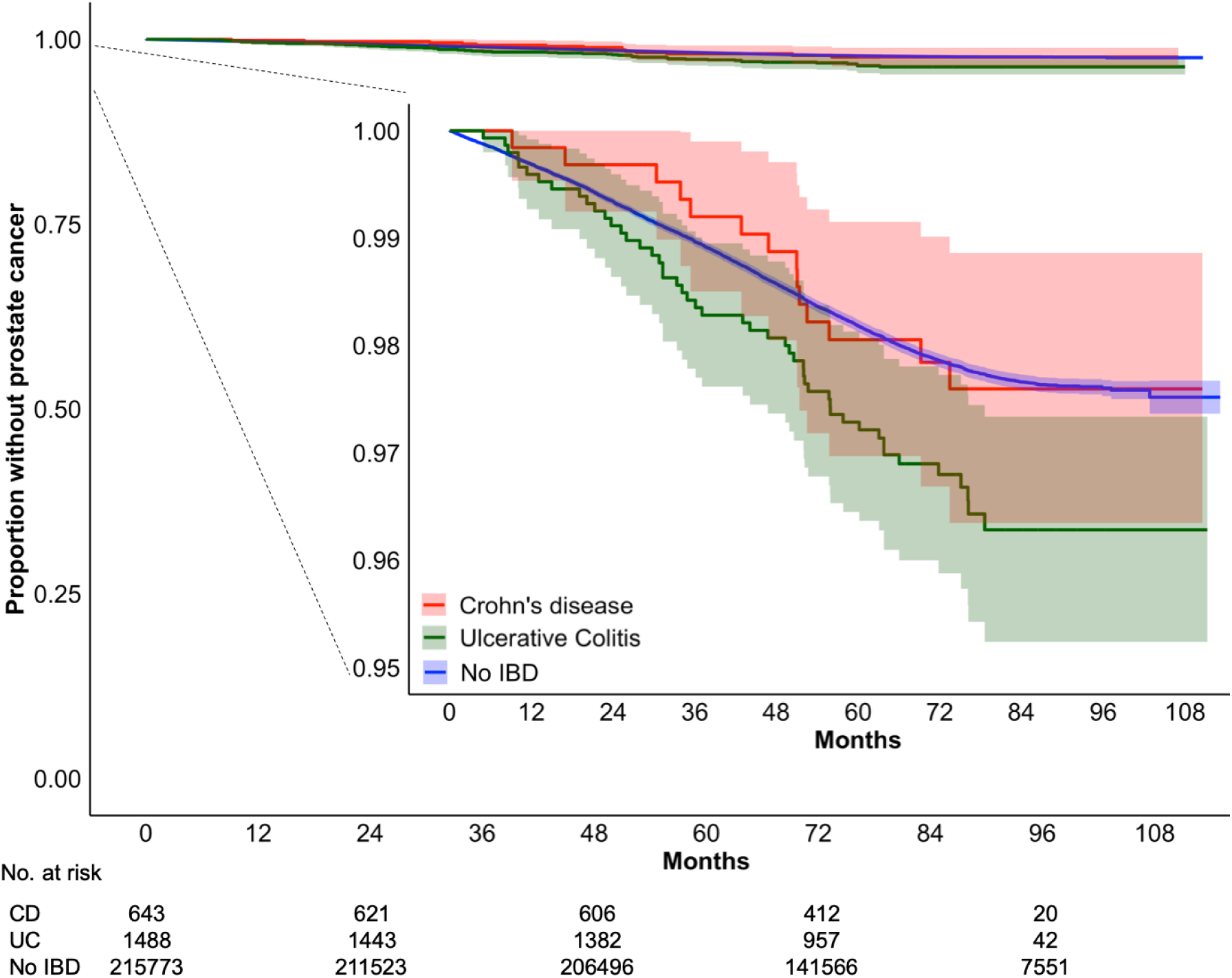
Kaplan-Meier analysis of prostate cancer incidence by inflammatory bowel disease subtype. Shading represents 95% confidence intervals. Log-rank comparing CD to no IBD p=0.95 and UC to No IBD p=0.0023. Abbreviations: CD = Crohn’s disease; UC = Ulcerative colitis; IBD = Inflammatory bowel disease

## Discussion

IBD is a chronic inflammatory condition with a growing prevalence, affecting at least 0.3% of individuals in developed countries^21^. In a large, prospective cohort of men in the UK, where PC screening is low (∼27% in our cohort), we found a positive association between IBD and PC. This association was driven by an approximate 50% increase in PC risk among men with UC. These findings suggest that even outside the setting of routine PC screening, men with IBD are at an increased risk of PC.

The limited research into IBD and PC has demonstrated conflicting results. In a recent, large, retrospective study at a single medical center, men with IBD undergoing PC screening had a greater than four-fold increase in incident PC and high-grade PC compared to men without IBD^9^. In a case-control study within a shared, equal-access healthcare system, men with IBD had a 70% increased risk of PC^7^. In contrast, other studies and a meta-analysis found no clear association between IBD and PC^14–16^. Three studies reported the association for UC but not CD^8,12,13^, as we observed in the present study. A pair of studies observed the association for both CD^10^ and UC^11^, although the associations were attenuated when cancers diagnosed within the first year after IBD diagnosis were excluded. Much of this prior research included men younger than 50 and thus at low risk of PC^8,12,13,15,16^, which may explain why two of these studies reported no IBD-PC association^15,16^. In contrast, the UK Biobank was designed to study age-related diseases^20^.

Several mechanisms may explain the potential relationship between IBD and PC. Chronic inflammation is a risk factor for cancer development in various solid tumors and may contribute to prostate tumorigenesis by inducing DNA damage and promoting carcinogenic epigenetic alterations^22^. Chronic inflammation may play a role in the well-established association between IBD and colorectal cancer^6^, though it is unknown whether chronic gut inflammation leads to changes in the prostatic inflammatory milieu. Transmission of gut inflammation to the prostate may occur via local inflammation in the rectum, which is nearly universal in UC and less frequently observed in CD. These IBD subtypes have distinct clinical and pathologic features^23^. While local inflammation may help to explain our observation and prior reports that UC, not CD, is associated with PC, further research is warranted, in particular the documentation of adjacent rectal inflammation in patients with UC vs. CD. An alternative factor may be elevated serum (systemic) inflammatory markers, which are known to occur in IBD^24^ and may play a role in PC development and progression^22^. In contrast, immunomodulatory medications commonly used in IBD have been associated with other extra-intestinal malignancies^14,25^, suggesting immunosuppression may also be responsible for prostate tumorigenesis or progression. Lastly, shared underlying genetics may explain the IBD-PC association, although preliminary evidence across common gene variants has not detected genetic correlations for either UC or CD with PC^26^.

The potential link between IBD and PC has important implications for screening and detection of PC. While recommendations are controversial, some guideline panels have supported more aggressive screening in high risk populations^4,5^. Older age, African-ancestry, and family history of PC have been consistently identified as risk factors for PC development^27^. Our study suggests that IBD may be an independent risk factor for PC, but future study is needed to determine how to appropriately apply this finding to patient screening practices.

There are several important limitations of our study. First, we were unable to account for frequency of healthcare encounters in our analysis of incident PC. Men with IBD have more frequent healthcare encounters^28^, are more likely to undergo rectal examinations, and may be subject to opportunistic screening. Our analysis was adjusted for whether the participant had a PSA test prior to entry into the UK Biobank, however, we were unable to account for number of prior PSA tests or digital rectal examinations. Nevertheless, in a sensitivity analysis, we observed that in men with no history of a PSA test, the hazard ratios were greater by 10% and 12% for IBD and UC, respectively compared to the models including all men and with PSA test as a covariate. Second, the release of the UK Biobank data at the time we performed the analysis did not provide information regarding cancer grade or stage; thus, we were unable to differentiate between low-risk and clinically significant cancer. Third, given the relatively few cancer diagnoses overall, we report incidence of PC diagnosis, but not morbidity or mortality related to diagnosis and treatment. Lastly, important characteristics of the UK Biobank limit the generalizability of these results. Participants in the study, compared to nonparticipants in the UK, are noted to be older, of higher socioeconomic status, predominantly Caucasian, and generally healthy—all of which have important relevance in PC screening and diagnosis. In addition, participants of the UK Biobank have lower all-cause mortality and total cancer incidence compared to nonparticipants which may suggest that our results under-estimate the burden of PC in men with IBD in the UK^29^.

## Conclusions

In this large-scale cohort study outside the setting of widespread PC screening, men with IBD had an increased risk of incident PC compared to men without IBD. Future work is needed to validate this association accounting for PC screening and other covariates, and determine potential mechanisms of prostate tumorigenesis in men with IBD. Ultimately, this work could provide an avenue for incorporating information about IBD into screening decisions for PC.

## Data Availability

UK Biobank is an open access resource. Bona fide researchers can apply to use the UK Biobank dataset by registering and applying at https://www.ukbiobank.ac.uk/register-apply/. All results presented in this manuscript, including the code used to generate them, will be returned to UK Biobank within 6 mo of publication at which point they are made available for researchers to request (subject to UK Biobank approval).

## Acknowledgements

This research was funded by the following NIH grants: NCI R25CA112355, R01CA201358, and NIA T32AG049663. This research was conducted with approved access to UK Biobank data under application number 14105.

## References

1. Bray F, Ferlay J, Soerjomataram I, Siegel RL, Torre LA, Jemal A. Global cancer statistics 2018: GLOBOCAN estimates of incidence and mortality worldwide for 36 cancers in 185 countries. CA Cancer J Clin. 2018;68(6):394–424. doi:10.3322/caac.21492

2. Hugosson J, Roobol MJ, Månsson M, et al. A 16-yr Follow-up of the European Randomized study of Screening for Prostate Cancer. Eur Urol. 2019;76(1):43–51. doi:10.1016/j.eururo.2019.02.009

3. Loeb S, Bjurlin MA, Nicholson J, et al. Overdiagnosis and overtreatment of prostate cancer. Eur Urol. 2014;65(6):1046–1055. doi:10.1016/j.eururo.2013.12.062

4. US Preventive Services Task Force, Grossman DC, Curry SJ, et al. Screening for Prostate Cancer: US Preventive Services Task Force Recommendation Statement. JAMA. 2018;319(18):1901–1913. doi:10.1001/jama.2018.3710

5. Gandaglia G, Albers P, Abrahamsson P-A, et al. Structured Population-based Prostate-specific Antigen Screening for Prostate Cancer: The European Association of Urology Position in 2019. Eur Urol. 2019;76(2):142–150. doi:10.1016/j.eururo.2019.04.033

6. Dulai PS, Sandborn WJ, Gupta S. Colorectal Cancer and Dysplasia in Inflammatory Bowel Disease: A Review of Disease Epidemiology, Pathophysiology, and Management. Cancer Prev Res (Phila Pa). September 2016. doi:10.1158/1940-6207.CAPR-16-0124

7. Mosher CA, Brown GR, Weideman RA, et al. Incidence of Colorectal Cancer and Extracolonic Cancers in Veteran Patients With Inflammatory Bowel Disease. Inflamm Bowel Dis. 2018;24(3):617–623. doi:10.1093/ibd/izx046

8. Kappelman MD, Farkas DK, Long MD, et al. Risk of Cancer in Patients with Inflammatory Bowel Diseases: a Nationwide Population-Based Cohort Study with 30 Years of Follow Up. Clin Gastroenterol Hepatol Off Clin Pract J Am Gastroenterol Assoc. 2014;12(2):265-73.e1. doi:10.1016/j.cgh.2013.03.034

9. Burns JA, Weiner AB, Catalona WJ, et al. Inflammatory Bowel Disease and the Risk of Prostate Cancer. Eur Urol. 2019;75(5):846–852. doi:10.1016/j.eururo.2018.11.039

10. Hemminki K, Li X, Sundquist J, Sundquist K. Cancer risks in Crohn disease patients. Ann Oncol Off J Eur Soc Med Oncol. 2009;20(3):574–580. doi:10.1093/annonc/mdn595

11. Hemminki K, Li X, Sundquist J, Sundquist K. Cancer risks in ulcerative colitis patients. Int J Cancer. 2008;123(6):1417–1421. doi:10.1002/ijc.23666

12. Jess T, Horváth-Puhó E, Fallingborg J, Rasmussen HH, Jacobsen BA. Cancer risk in inflammatory bowel disease according to patient phenotype and treatment: a Danish population-based cohort study. Am J Gastroenterol. 2013;108(12):1869–1876. doi:10.1038/ajg.2013.249

13. Jung YS, Han M, Park S, Kim WH, Cheon JH. Cancer Risk in the Early Stages of Inflammatory Bowel Disease in Korean Patients: A Nationwide Population-based Study. J Crohns Colitis. 2017;11(8):954–962. doi:10.1093/ecco-jcc/jjx040

14. Pedersen N, Duricova D, Elkjaer M, Gamborg M, Munkholm P, Jess T. Risk of extra-intestinal cancer in inflammatory bowel disease: meta-analysis of population-based cohort studies. Am J Gastroenterol. 2010;105(7):1480–1487. doi:10.1038/ajg.2009.760

15. Wilson JC, Furlano RI, Jick SS, Meier CR. A population-based study examining the risk of malignancy in patients diagnosed with inflammatory bowel disease. J Gastroenterol. 2016;51(11):1050–1062. doi:10.1007/s00535-016-1199-8

16. van den Heuvel TRA, Wintjens DSJ, Jeuring SFG, et al. Inflammatory bowel disease, cancer and medication: Cancer risk in the Dutch population-based IBDSL cohort. Int J Cancer. 2016;139(6):1270–1280. doi:10.1002/ijc.30183

17. Sudlow C, Gallacher J, Allen N, et al. UK biobank: an open access resource for identifying the causes of a wide range of complex diseases of middle and old age. PLoS Med. 2015;12(3):e1001779. doi:10.1371/journal.pmed.1001779

18. Williams N, Hughes LJ, Turner EL, et al. Prostate-specific antigen testing rates remain low in UK general practice: a cross-sectional study in six English cities. BJU Int. 2011;108(9):1402–1408. doi:10.1111/j.1464-410X.2011.10163.x

19. Prostate Cancer. https://legacyscreening.phe.org.uk/prostatecancer. Accessed December 23, 2019.

20. Key documents | UK Biobank. https://www.ukbiobank.ac.uk/key-documents/. Accessed November 19, 2019.

21. Ng SC, Shi HY, Hamidi N, et al. Worldwide incidence and prevalence of inflammatory bowel disease in the 21st century: a systematic review of population-based studies. Lancet Lond Engl. 2018;390(10114):2769–2778. doi:10.1016/S0140-6736(17)32448-0

22. Sfanos KS, Yegnasubramanian S, Nelson WG, De Marzo AM. The inflammatory microenvironment and microbiome in prostate cancer development. Nat Rev Urol. 2018;15(1):11–24. doi:10.1038/nrurol.2017.167

23. Souza HSP de, Fiocchi C. Immunopathogenesis of IBD: current state of the art. Nat Rev Gastroenterol Hepatol. 2016;13(1):13–27. doi:10.1038/nrgastro.2015.186

24. Vrakas S, Mountzouris KC, Michalopoulos G, et al. Intestinal Bacteria Composition and Translocation of Bacteria in Inflammatory Bowel Disease. PloS One. 2017;12(1):e0170034. doi:10.1371/journal.pone.0170034

25. Biancone L, Onali S, Petruzziello C, Calabrese E, Pallone F. Cancer and immunomodulators in inflammatory bowel diseases. Inflamm Bowel Dis. 2015;21(3):674–698. doi:10.1097/MIB.0000000000000243

26. Lindström S, Finucane H, Bulik-Sullivan B, et al. Quantifying the genetic correlation between multiple cancer types. Cancer Epidemiol Biomark Prev Publ Am Assoc Cancer Res Cosponsored Am Soc Prev Oncol. 2017;26(9):1427–1435. doi:10.1158/1055-9965.EPI-17-0211

27. Pernar CH, Ebot EM, Wilson KM, Mucci LA. The Epidemiology of Prostate Cancer. Cold Spring Harb Perspect Med. 2018;8(12):a030361. doi:10.1101/cshperspect.a030361

28. Park KT, Ehrlich OG, Allen JI, et al. The Cost of Inflammatory Bowel Disease: An Initiative From the Crohn’s & Colitis Foundation. Inflamm Bowel Dis. May 2019. doi:10.1093/ibd/izz104

29. Fry A, Littlejohns TJ, Sudlow C, et al. Comparison of Sociodemographic and Health-Related Characteristics of UK Biobank Participants With Those of the General Population. Am J Epidemiol. 2017;186(9):1026–1034. doi:10.1093/aje/kwx246

